# Interactive Segmentation of Lung Tissue and Lung Excursion in Thoracic Dynamic MRI Based on Shape-guided Convolutional Neural Networks

**DOI:** 10.1101/2024.05.03.24306808

**Authors:** Lipeng Xie, Jayaram K. Udupa, Yubing Tong, Joseph M. McDonough, Patrick J. Cahill, Jason B. Anari, Drew A. Torigian

## Abstract

**Purpose:** Lung tissue and lung excursion segmentation in thoracic dynamic magnetic resonance imaging (dMRI) is a critical step for quantitative analysis of thoracic structure and function in patients with respiratory disorders such as Thoracic Insufficiency Syndrome (TIS). However, the complex variability of intensity and shape of anatomical structures and the low contrast between the lung and surrounding tissue in MR images seriously hamper the accuracy and robustness of automatic segmentation methods. In this paper, we develop an interactive deep-learning based segmentation system to solve this problem.

**Material & Methods:** Considering the significant difference in lung morphological characteristics between normal subjects and TIS subjects, we utilized two independent data sets of normal subjects and TIS subjects to train and test our model. 202 dMRI scans from 101 normal pediatric subjects and 92 dMRI scans from 46 TIS pediatric subjects were acquired for this study and were randomly divided into training, validation, and test sets by an approximate ratio of 5:1:4. First, we designed an interactive region of interest (ROI) strategy to detect the lung ROI in dMRI for accelerating the training speed and reducing the negative influence of tissue located far away from the lung on lung segmentation. Second, we utilized a modified 2D U-Net to segment the lung tissue in lung ROIs, in which the adjacent slices are utilized as the input data to take advantage of the spatial information of the lungs. Third, we extracted the lung shell from the lung segmentation results as the shape feature and inputted the lung ROIs with shape feature into another modified 2D U-Net to segment the lung excursion in dMRI. To evaluate the performance of our approach, we computed the Dice coefficient (DC) and max-mean Hausdorff distance (MM-HD) between manual and automatic segmentations. In addition, we utilized Coefficient of Variation (CV) to assess the variability of our method on repeated dMRI scans and the differences of lung tidal volumes computed from the manual and automatic segmentation results.

**Results:** The proposed system yielded mean Dice coefficients of 0.96±0.02 and 0.89±0.05 for lung segmentation in dMRI of normal subjects and TIS subjects, respectively, demonstrating excellent agreement with manual delineation results. The Coefficient of Variation and p-values show that the estimated lung tidal volumes of our approach are statistically indistinguishable from those derived by manual segmentations.

**Conclusions:** The proposed approach can be applied to lung tissue and lung excursion segmentation from dynamic MR images with high accuracy and efficiency. The proposed approach has the potential to be utilized in the assessment of patients with TIS via dMRI routinely.

## 1. Introduction

### Background

Thoracic insufficiency syndrome (TIS) is a condition that affects pediatric patients with spine and chest wall deformities, leading to respiratory impairment, which is characterized by both restrictive and, less commonly, obstructive lung disease due to changes in spine and rib configuration, which reduce lung volume, stiffen the chest wall, and weaken respiratory muscles [1, 2]. Collaborative efforts between pediatric pulmonologists and spine surgeons are necessary to determine the best treatment options, including non-surgical and surgical strategies, timing of surgery, and medical supportive care [3]. Management and treatment of patients with TIS involve growth-sparing surgery, such as the use of the Vertical Expandable Prosthetic Titanium Rib (VEPTR), to preserve vertical growth before a definitive spinal fusion at skeletal maturity [4, 5]. To quantitively analyze the changes in regional dynamic thoracic function before and after surgical correction of TIS, the clinical parameters of TIS, including forced vital capacity [6], thoracic and lumbar Cobb angles [7], and resting breathing rate (RR) [8], etc. are usually measured. However, these clinical parameters have not demonstrated significant changes of lung function after surgery, especially when regional components are concerned [9]. To address this problem, Tong et al. proposed the quantitative dynamic MRI (QdMRI) method for providing valuable insights into the impact of surgery on lung volumes [9, 10], by utilizing measurements of lung volume at end-inspiration and end-expiration, lung tidal volume, chest wall excursion volume, and diaphragm excursion volume separately for each hemi-chest. In addition, QdMRI is more suitable for analyzing the properties of multiple objects of interest beyond the lungs for very young TIS patients compared with the ultrashort echo time (UTE) MRI [11, 12].

Thoracic dynamic magnetic resonance imaging (dMRI) has been used to assess regional thoracic function in patients with TIS [9], to evaluate lung function in adolescent idiopathic scoliosis (AIS) [13], and to analyze diaphragmatic motion [14]. In such applications, lung tissue and lung excursion segmentation in dMRI plays a crucial role in quantitative analysis. However, manual delineation of the lung tissue and lung excursion is exceedingly time-consuming and requires significant labor. Furthermore, this approach is susceptible to inconsistencies between different individuals. Consequently, there is a pressing requirement for medical professionals and researchers to possess a robust, efficient, accurate, and precise technique for segmenting the lung tissue and excursion regions in order to enhance productivity workflow. Nevertheless, development of robust systems for this purpose is challenging due to several factors: i) complex variability of grayscale intensity among different dMRI acquisition protocols, ii) complex structure and variability of lung shape across different patients, especially in TIS, iii) low image contrast and weak boundary between lung tissue and surrounding tissues, and iv) often inadequate signal-to-noise ratio on MR images. In addition, the shortage of high-quality annotation data seriously limits the development and evaluation of advanced lung segmentation approaches.

### Related work

To solve the above problems, there have been some published studies on lung segmentation in MRI and computed tomography (CT), which can be divided into unsupervised and supervised models [15]. The unsupervised approaches are based on unsupervised machine learning (ML) models and traditional image segmentation methods without the requirement of data annotation. In [16], an active contour model was developed that provides a smooth boundary and accurately captures the high curvature features of the lungs from MR images. In [17], atlas-based methods were used to segment the lungs in multi-sequence proton MR images. Hassani et al. [18] proposed an automated hybrid method for lung segmentation based on both mathematical morphology and the region growing algorithm, where seed points are selected automatically without any user interaction. Similarly, Mansoor et al. [19] presented a novel method for pathological lung segmentation in CT using a fuzzy connectedness (FC) algorithm and texture-based features, in which the seed is automatically selected for initial FC segmentation based on voxel intensity and geometrical knowledge. Furthermore, Tong et al. proposed an interactive iterative relative fuzzy connectedness approach for lung segmentation on thoracic 4D dynamic MR images, achieving a mean value of 0.91 and a standard deviation of 0.03 for true positive volume fraction [20]. Yet, the above methods have low computation efficiency and may be affected by noise in the MR and CT images. To improve robustness and efficiency, some researchers have utilized a supervised machine learning model to construct lung segmentation systems. For instance, Neil et al. [21] designed hybrid model, in which a hierarchical detection network (HDN) was used to detect stable landmarks on the surface of the lungs to robustly initialize a level-set model for delineating lung contours. In [22], an artificial neural network (ANN) was employed to segment the lung tissue in matrix pencil decomposition MRI, achieving a mean value of 0.89 and a standard deviation of 0.03 for Dice similarity coefficient (DSC).

Recently, deep learning (DL) has revolutionized medical image analysis and achieved superior performance, involving use of deep neural networks to learn hierarchical feature representations from medical images, and eliminating the need for hand-designed features [23]. Deep learning models have been successfully applied in various medical imaging applications and have shown better capabilities in segmenting and classifying medical images, such as MRI and CT images, compared to conventional image processing and machine learning techniques [24, 25]. Several DL-based lung segmentation approaches have been proposed. For instance, studies [26–28] utilized a convolutional neural network (CNN) to segment the lung tissue in CT images, obtaining excellent segmentation performance and high efficiency. Similarly, Astley et al. [29] constructed a ventilated lung segmentation system based on 3D CNN for multi-nuclear hyperpolarized gas MRI. The experimental data demonstrated that the DL-based lung segmentation approaches outperformed the unsupervised ML-based methods such as K-means and spatial fuzzy C-means. In their latest work [30], a proposed implementable DL segmentation algorithm was used to produce accurate lung segmentations on a large, multi-center, multi-acquisition, and multi-disease set of MRI scans. In [31], we presented an automatic DL-based lung segmentation system, including two CNNs: i) a corner-points detection network for locating the lung in MRI and extracting the region of interest (ROI) of lung, and ii) a lung segmentation network for pixel-wise classification in the lung ROI. The proposed method achieves a high mean value of 0.97 and a low standard deviation of 0.02 for 3D DSC, demonstrating that the lung ROI can improve lung segmentation performance. However, the lung ROI detection accuracy of our method was low for dMRI obtained in TIS patients, reducing the lung segmentation performance for TIS patients. In addition, the above methods cannot produce segmentations of the excursion regions of the chest wall and diaphragm in dMRI and meet the requirement of lung tidal volume measurement.

In this paper, we propose a semi-automated and easy-to-use lung segmentation system for use in TIS patients (often with severe deformities of the chest) based on interactive lung ROI strategy and deep CNN for dMRI, achieving a high agreement with reference standard manual segmentations. The main innovations of this study include: 1) a semi-automatic, robust, and accurate segmentation system for lung tissue and lung excursion in dMRI; 2) an accurate and adaptive interactive lung ROI detection strategy based on the corner-points concept and linear interpolation; 3) a novel design of network architecture using the hand-crafted lung shape feature to segment the chest wall and diaphragm excursion regions from lung tissue; 4) demonstration of as close to the highest possible lung segmentation performance considering the quality of the lung ROI in both normal subjects and TIS patients; 5) demonstration of high agreement between manually and automatically measured lung tidal volumes; and 6) comparison with other methods from the literature to elucidate the comprehensive advantages of our approach for application in patients with TIS.

The following sections are arranged as follows. The details about our method are introduced in Section 2 including preparatory operations, interactive ROI strategy, segmentation network design, post-processing method, and data augmentation. In Section 3, we illustrate the qualitative and quantitative results including lung ROI detection, lung segmentation, lung excursion segmentation, estimated tidal volumes, and comparisons between our approach and related studies. Finally, we summarize our conclusions in Section 4.

A preliminary report of this work appeared in the proceedings of the 2022 SPIE Medical Imaging Conference [31]. The present paper includes the following enhancements over the conference paper: (i) a detailed and comprehensive literature review about lung segmentation in MRI; (ii) a full description of our approach including dMRI image acquisition, interactive ROI strategy, ROI-guided lung segmentation model, and shape-guided lung excursion segmentation model; and (iii) a detailed demonstration of our experimental results on a much larger data set including illustrative segmentation examples, quantitative evaluation metric values, lung tidal volume computation results, and comparisons with other approaches in the literature.

## 2. Materials & Methods

### 2.1 Method overview

In our study, we focus on the segmentation of lung tissue and lung excursion region from dMRI images. As illustrated in Figure 1, the lung region would expand with the diaphragm contracting and moving downward during the inspiration cycle. The movement of lung between the end-expiration and end-inspiration produces the lung excursion region, which can be used to estimate the respective tidal volumes. Thus, we firstly design an interactive segmentation model to obtain the lung region at end-expiration and end-inspiration and then construct the automatic lung excursion segmentation model based on the lung region segmentation.

**Figure 1.**
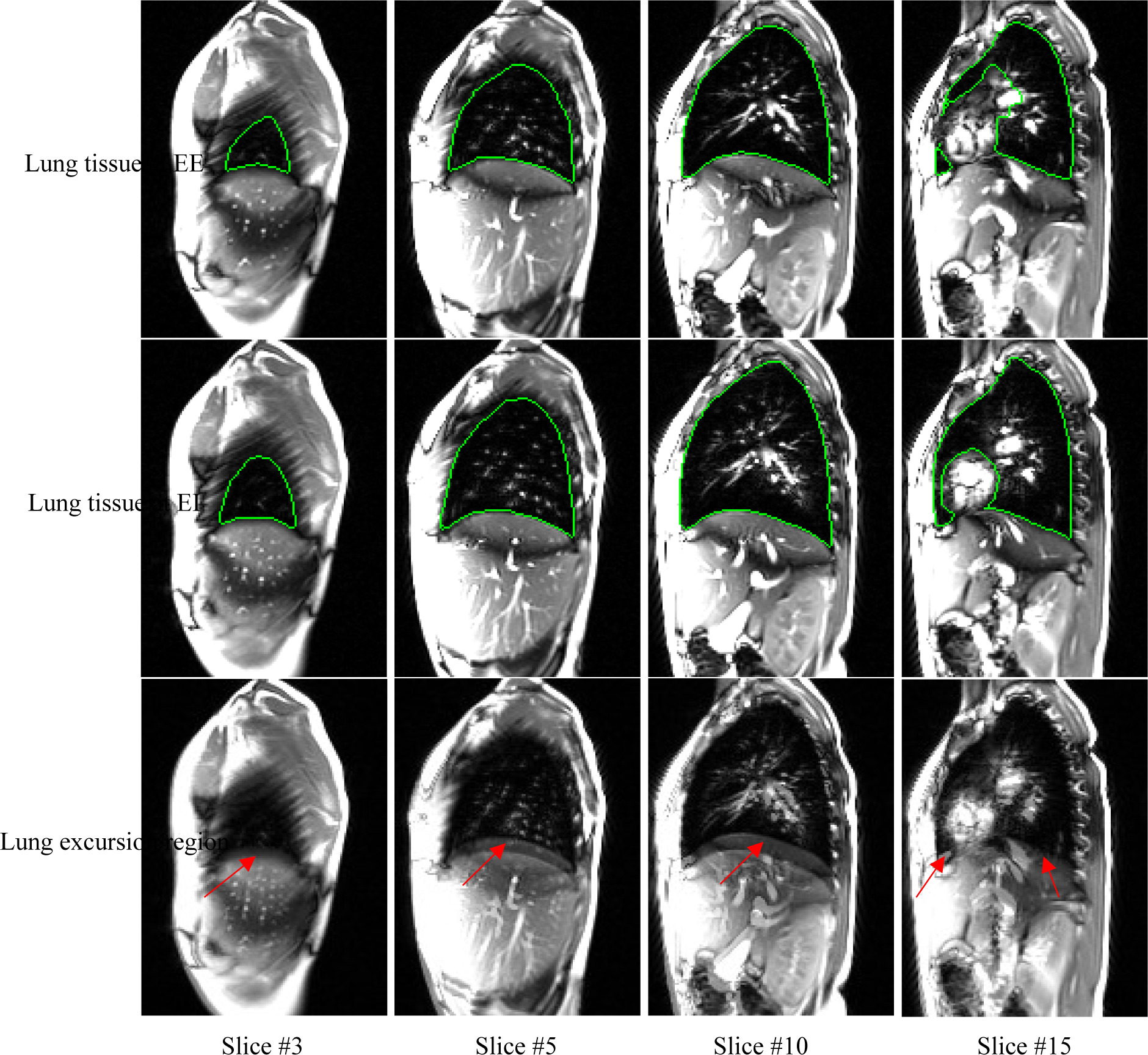
Illustration of lung tissue and lung excursion region in dMRI. 1^st^ - 2^nd^ rows: Lung tissue at end-expiration and end-inspiration (green boundaries). 3^rd^: lung excursion region (red arrows). 1^st^-4^th^ columns: From the right lung of one normal subject.

As shown in Figure 2, our study contains three main stages. The preparatory stage, in which we acquired and annotated thoracic dynamic 3D MRI scans and repeat-scans, divided into four parts: data sets for training, validation, testing, and volume variability study for evaluating the performance of our system. The model-building stage involves the following steps: i) Designing the interactive ROI strategy for localizing lungs based on corner-points of the bounding box and a linear interpolating method. ii) Developing a CNN based segmentation model for the lung guided by the ROI of the lung from the previous step. iii) Constructing the segmentation network for the excursion region based on U-Net and shape-prior information. Excursion region refers to the region excursed by each hemi-chest wall and hemi-diaphragm, which is required for estimating the respective tidal volumes. iv) Devising a post-processing method for reducing the false positive rate of excursion region. v) Performing data augmentation and DL model training. vi) The evaluation stage employed the trained models to segment dMRIs in the testing and repeat-scan sets.

**Figure 2.**
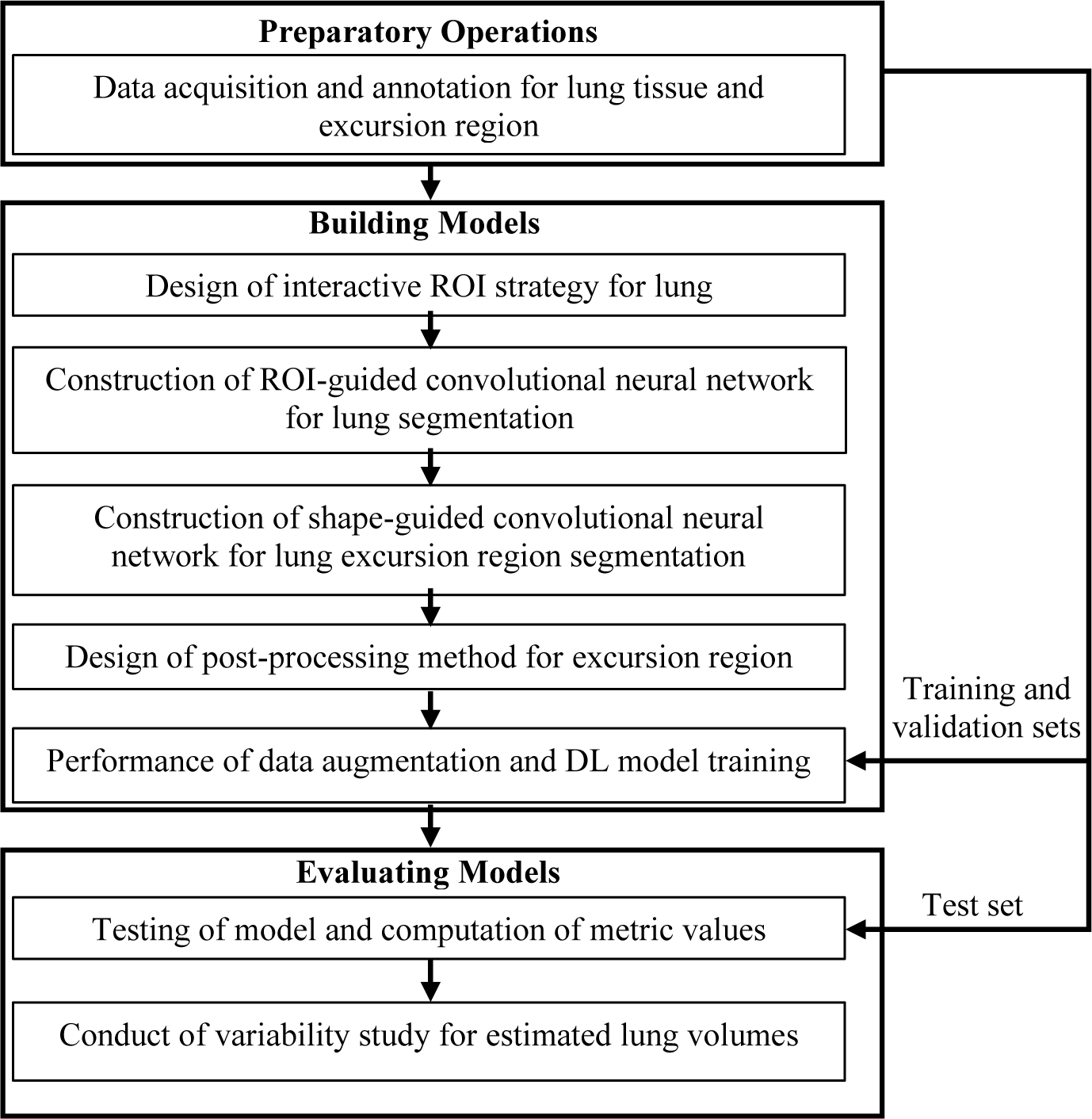
Processing pipeline depicting the main stages in our study.

### 2.2 Preparatory operations

a. Acquisition and annotation of datasets

All dynamic MRI studies were acquired under an IRB-approved research protocol at the Children’s Hospital of Philadelphia along with Health Insurance Portability and Accountability Act waiver.

Our dynamic MRI protocol was as follows: 3.0 T MRI scanner (Siemens Healthcare, Erlangen, Germany), true-FISP bright-blood imaging with steady-state precession sequence; TR = 3.82 ms, TE = 1.91 ms, voxel size ∼1×1×6 mm^3^; 320 × 320 × 38 matrix; bandwidth, 558 Hz; flip angle, 76°. For each sagittal location through the thorax, slice data were gathered over 8–14 tidal breathing cycles at about 480 ms per slice. The acquisition time was ∼45 minutes. The number of 2D slices acquired in this manner was typically 2000 to 3000. From these data, we then constructed a 4D image comprising of 200-300 slices using a previously published method [32, 33]. The 4D image represents the full 3D dynamic chest over one respiratory cycle comprising of 5-8 respiratory time points.

To analyze the difference in lung tissue and lung excursion region segmentation performance between normal subjects and TIS patients for our method, we collected dMRI scans from a large number of normal subjects and TIS patients and separated the datasets into two sub sets, as summarized in Table 1. For segmentation tasks, we focused on the 3D images corresponding to the end-inspiration (EI) phase and the end-expiration (EE) phase of the 4D image. The “Normal” sub dataset was from 101 normal subjects, including 202 3D frames with 5972 2D slices, while the TIS sub dataset was from 46 TIS patients, including 92 3D frames with 3676 2D slices. To train and evaluate our model, we divided the dMRI scans into training, validation, and testing data sets by an approximate ratio of 5:1:4. To evaluate the variability of estimated lung volume in repeated scans, we performed repeat scans of 10 normal subjects by performing a second scan after a short break at the end of the 1^st^ scan. In each of the first and the second scan, there were 60 3D frames in total (roughly 6 frames covering different phases of the respiratory cycle) which were utilized to test the repeatability of segmentation; see Table 2.

**Table 1.** Summary of available dynamic MRI scans for training and testing the segmentation models.

|  |  |  |  |  |
| --- | --- | --- | --- | --- |
| Dataset | Normal subjects |  | TIS subjects |  |
|  | Subjects | 3D frames | Subjects | 3D frames |
| Training set | 50 | 100 (2938 slices) | 22 | 44 (1732 slices) |
| Validation set | 11 | 22 (642 slices) | 4 | 8 (324 slices) |
| Testing set | 40 | 80 (2392 slices) | 20 | 40 (1620 slices) |
| Total | 101 | 202 (5972 slices) | 46 | 92 (3676 slices) |
| Voxel size (mm<sup>3</sup>) | 0.98x0.98x6.00~1.46x1.46x6.00 |  |  |  |

**Table 2.**
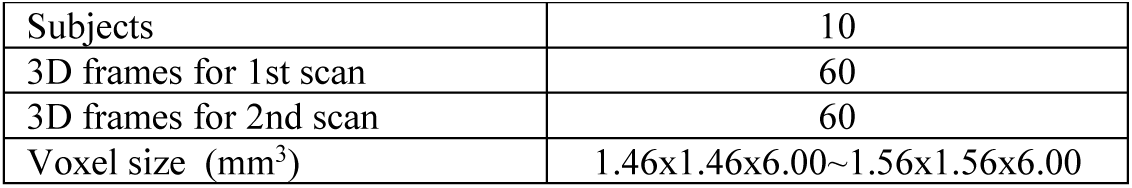
Summary of available dynamic dMRI scans using the repeat scan acquisition protocol for estimating variability.

|  |  |
| --- | --- |
| Subjects | 10 |
| 3D frames for 1st scan | 60 |
| 3D frames for 2nd scan | 60 |
| Voxel size (mm<sup>3</sup>) | 1.46x1.46x6.00~1.56x1.56x6.00 |

The 4D constructed dMRI images listed in Table 1 were manually delineated using the open-source software CAVASS [34] for the lung tissue and lung excursion region by several rigorously trained medical interns and technicians with an appropriate training in human anatomy and the radiological appearance of the relevant structures. To ensure annotation quality, the training, guidance of the annotation process, and checking for quality were directed by a radiologist (DAT) with over 25 years of experience in MRI and thoracic radiology.

Data pre-processing method

To reduce the intensity variability due to different acquisition parameters, we employed an MRI signal intensity standardization method to pre-process the dMRI images. In addition, we utilized an image contrast enhancement method for dMRI images based on the min-max normalization model and lung intensity statistics as follows:

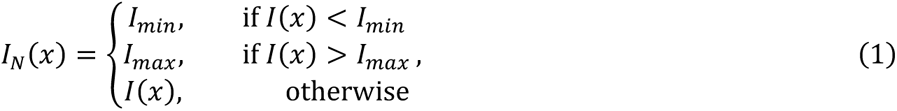

where *I*(*x*) and *I*_*N*_(*x*) represent the intensity of pixel *x* in the original image *I* and the pre-processed image *I*_*N*_, respectively, and *I*_*min*_and *I*_*max*_ represent the minimum and maximum intensity of lung tissue in MRI, respectively. This mapping is used to increase the contrast between the lung and surrounding tissues. In our experiments, we set the *I*_*min*_and *I*_*max*_ to 0 and 400, respectively, based on the statistical information from the training data set. To accelerate the convergence speed for the deep learning models, we rescaled the intensity range of *I*_*N*_(*x*) into [0, 1] as follows:

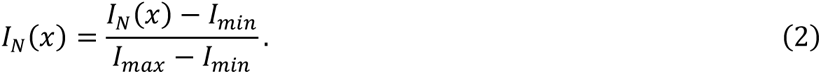

### 2.3 Interactive ROI strategy

In our previous work [31], we developed an automatic lung ROI strategy based on the corner-points detection network to improve lung segmentation performance, which can locate the lung tissue in dMRI scans of normal subjects using the bounding-box slice by slice. However, the lung detection error becomes large for TIS patients due to the deformed shape of the lung and its variability among slices. To ensure the ROI detection accuracy is sufficiently high for normal subjects and TIS patients, we design a novel interactive ROI strategy based on the corner-points concept and linear interpolation, as shown in Figure 3. The main steps in this method are as follows:

**Figure 3.**
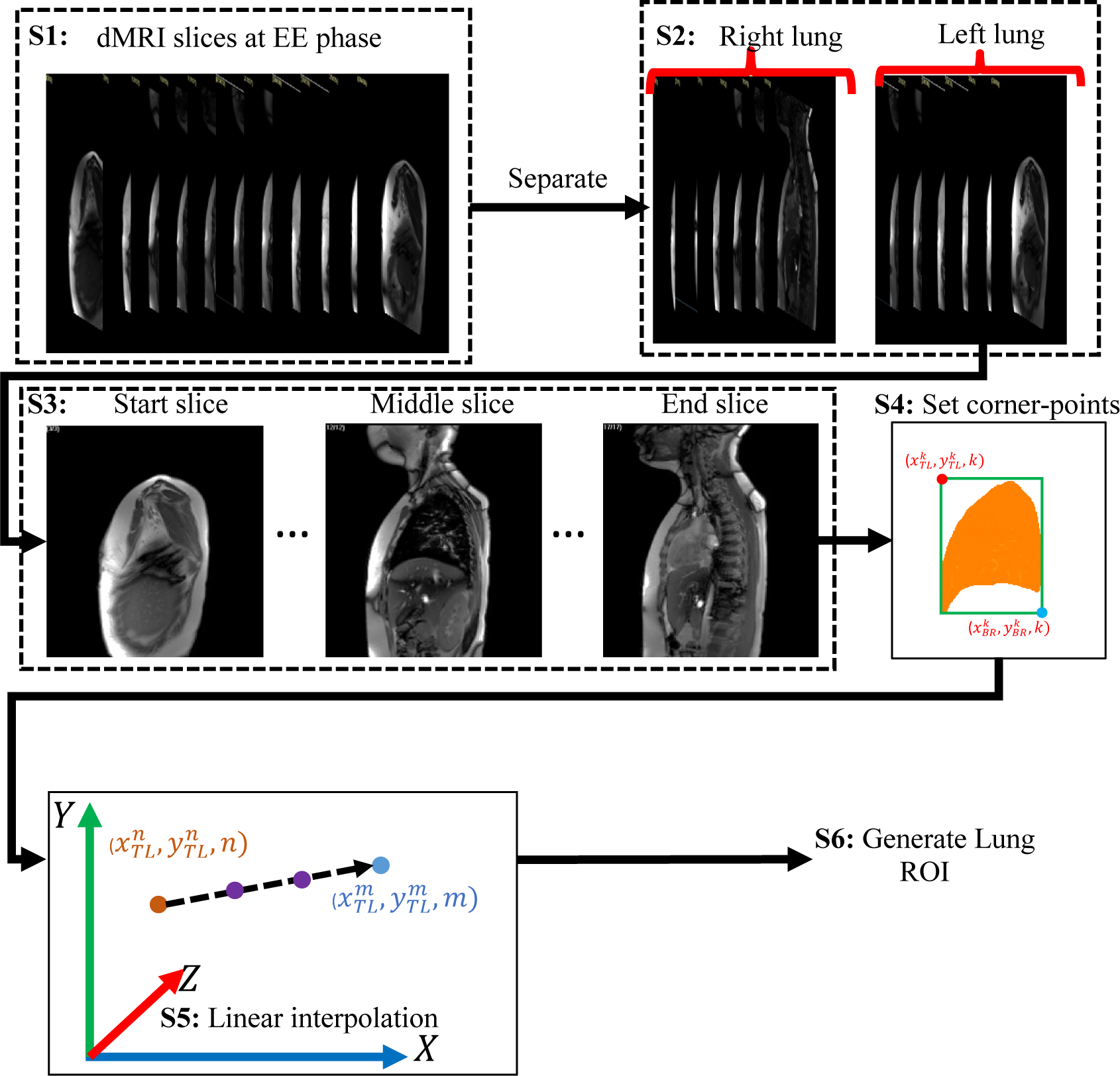
Illustration of the interactive ROI strategy.

S1: Identification of the 3D frame at the EE phase. Due to the high similarity among 3D frames in one dMRI scan, the same lung ROI found in one 3D frame can be applied in other 3D frames. In our method, we utilize the interactive ROI strategy to process the 3D frame at the EE phase.

S2: Separation of the right and left lungs. The 4D dMRI sequence was manually separated into two right lung and left lung. We compute the ROIs in right and left lungs separately. Note that the slice range of the right lung can overlap with that of the left lung due to disease-related deformations.

S3: Identification of key slices. Considering the dependence between adjacent slices, we select several key slices to conduct the interactive operations, including: 1) the start slice of the lung, which indicates the start position of the lung, 2) the end slice of the lung, which indicates the end position of the lung, and 3) the middle slice of the lung, which contains the maximum 2D lung tissue region. Note that the lung detection performance of the interactive ROI strategy is related to the number of key slices. Thus, the user can add more key slices between the start slice and middle slice or end slice to improve the ROI accuracy.

S4: Annotation of the corner-points. Our method requires the user to annotate the top-left (*x*_*TL*_, *y*_*TL*_) and bottom-right (*x*_*BR*_, *y*_*BR*_) points of ROI in the selected key slices. In general, the bounding box determined by the top-left and bottom-right points should cover the lung tissue.

S5: Interpolation of the coordinates for the interval corner-points. Given the coordinates of the top-left points (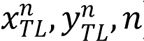) and (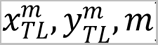) in the *n*-th and *m*-th slices, the coordinates of the top-left points between the *n*-th and *m*-th slices are computed by linear interpolation along the 3^rd^ dimension. The same operations are performed for the bottom-right points.

S6: Generation of the lung ROI. We utilize the coordinates of the corner-points in each slice to generate a rectangular ROI for the slice.

### 2.4 ROI-guided lung tissue segmentation network

Our joint interactive and CNN strategy is shown in Figure 4. Firstly, we utilized the interactive ROI strategy to generate the bounding-box for the lung tissue in 2D MRI slices. Then, the proposed image contrast enhancement method was applied to increase the pixel intensity difference between the lung and surrounding tissues. The ROIs found in 2D MRI slices in the previous step form the input data for the lung segmentation model. Note that we enlarge the size of the ROI by 1.2 times to ensure that the generated ROI always covers the lung region. The adjacent slices of the target slice were also inputted into the segmentation network as the context information to increase lung segmentation accuracy. Finally, the output pixel-wise probability map of the segmentation model was transformed into a binary image using a threshold of 0.5. To keep the image size of segmentation results consistent with the original images, we utilized zero-padding to embed the segmentations in the full image outside of the ROI. In addition, we utilized morphological operations to fill small holes in the lung segmentation results.

**Figure 4.**
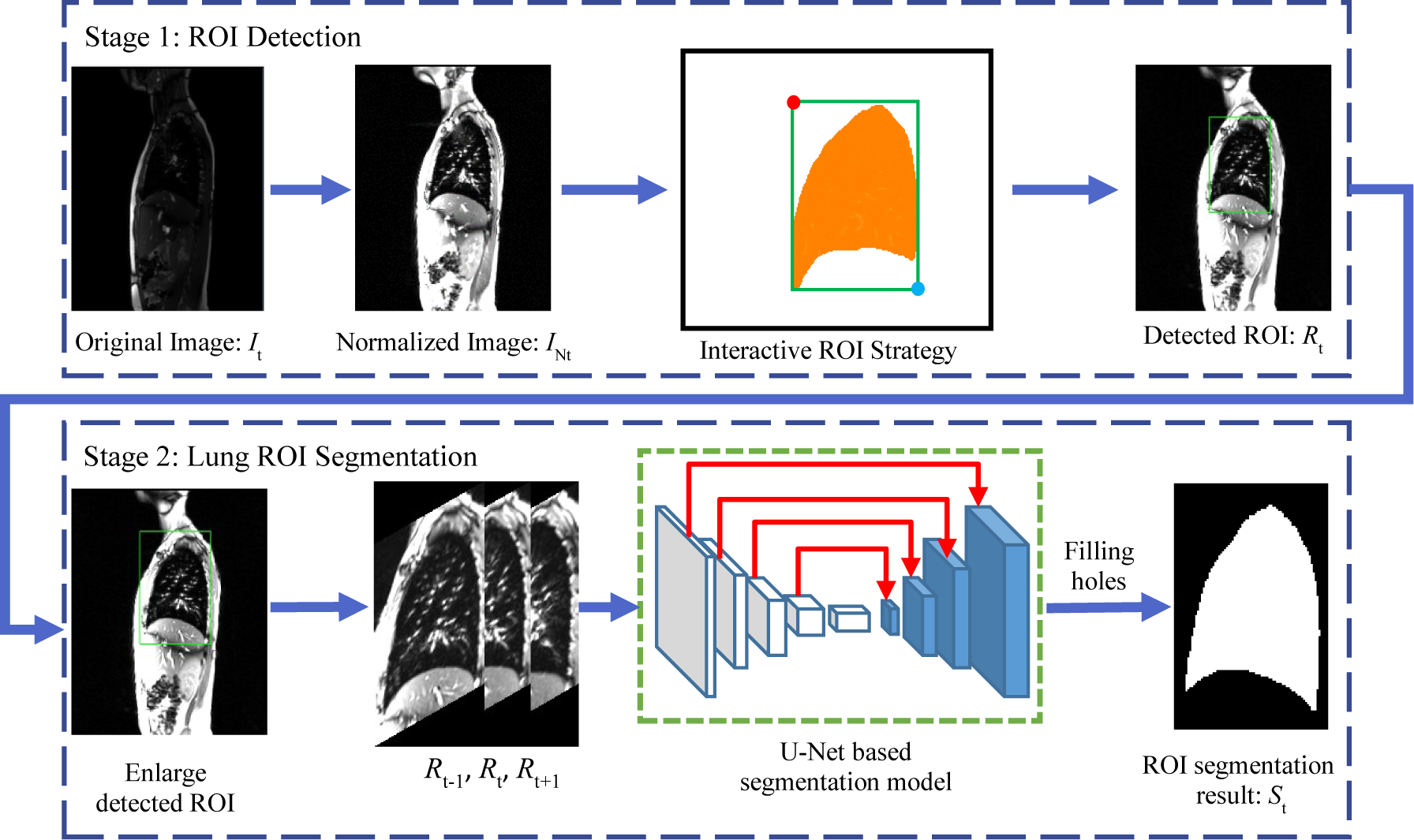
Illustration of ROI-guided U-Net for semi-automatic lung segmentation in dynamic MRI.

As shown in Figure 5, the architecture of the lung segmentation network includes three modules: i) encode network based on VGG19 [35] (left side) for learning the multi-scale features from the input images with size of 224×256x3, including 16 convolutional layers with 3×3 kernels, 4 max-pooling layers with stride 2, 3 dropout layers with rate 0.6, and 5 batch-normalization layers; ii) decode network based on FPN [36] (right side) for integrating the multi-scale features by linear interpolation and channel concatenation methods, including convolutional layers with 3×3 kernels, 4 up-sampling layers based on bilinear interpolation, and 4 concatenation layers; and iii) the pixel-wise classification module for computing the probability value of lung tissue pixel including 2 dropout layers with rate 0.6, 2 convolutional layers with 3×3 kernels, and 1 soft-max function. The main differences between our network and U-Net include replacing the de-convolutional layer with an up-sampling layer for reducing the number of parameters in our model and using the convolutional layer with 3×3 kernels and 32 output channels to reduce the redundancy of the feature fusion map.

**Figure 5.**
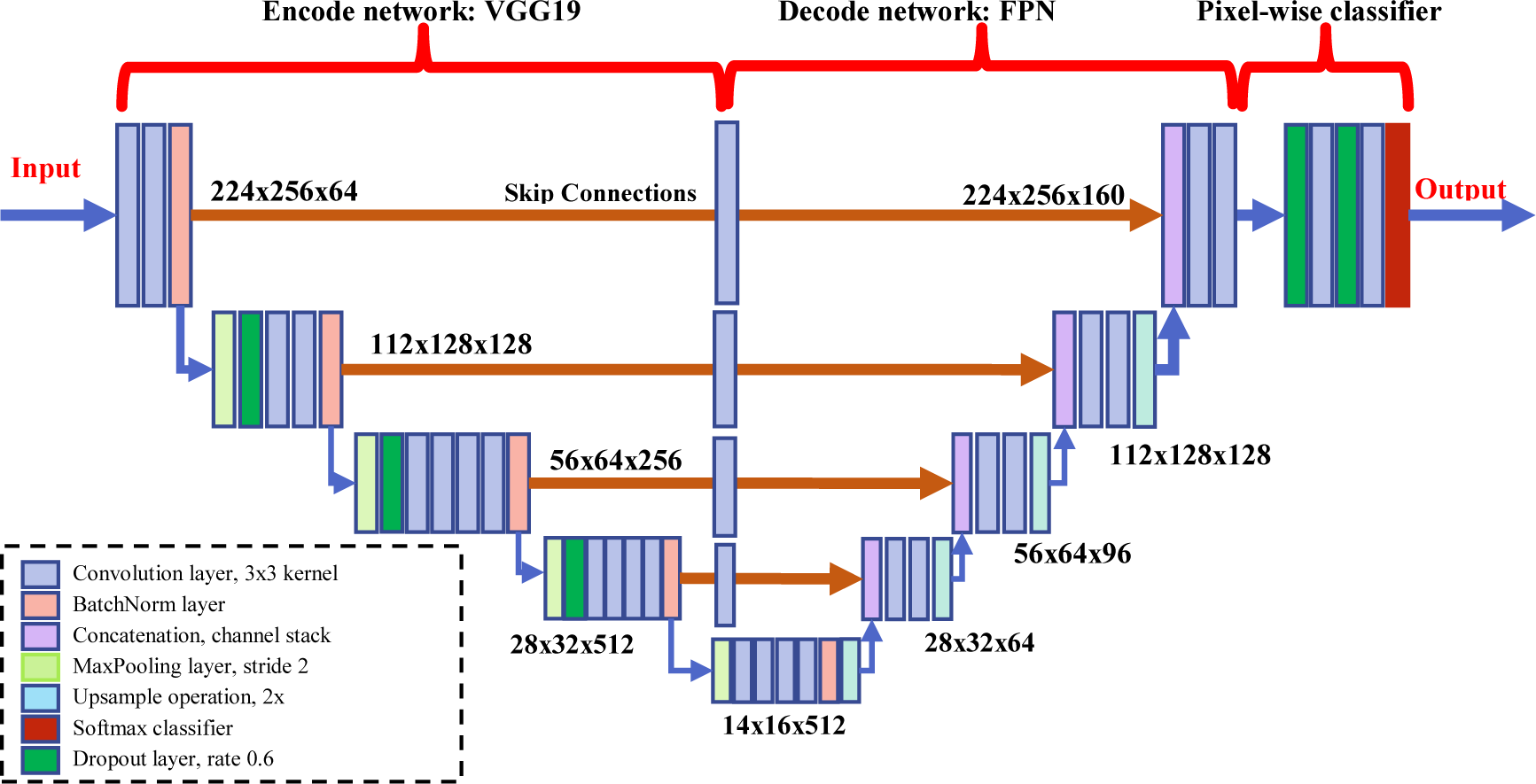
The architecture of lung segmentation network based on U-Net.

We utilized a loss function [37] that is a function of Dice, and explicitly FP and FN, to train the segmentation network:

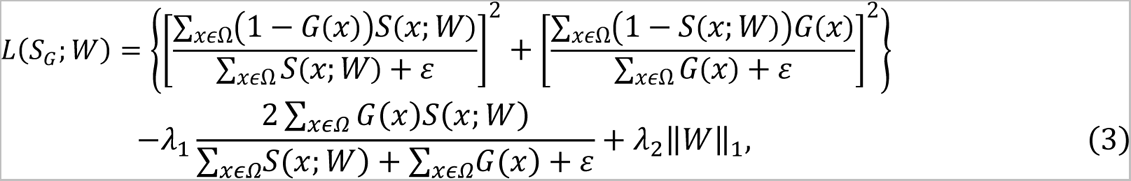

where *S*(*x*) denotes probabilistic value ranging from 0 to 1 output at pixel *x* for its classification, G(x) represents binary value at pixel *x*, *W* and Ω represent the parameters of the network and the image domain, respectively, ‖⋅‖_1_ is the L1-norm, and *λ*_1_ (> 0) and *λ*_2_ (> 0) serve as trade-off parameters among the three terms. The first two terms represent the false positive rate, false negative rate, and Dice index of the segmentation result. The 3^rd^ term was utilized as a sparsity constraint to reduce the over-fitting risk for our method.

### 2.5 Shape-guided lung excursion segmentation network

In this study, we propose a novel approach to segment the lung excursion region based on a CNN and shape prior-knowledge, as shown in Figure 6. In this method, we utilized the pixel-wise logical XOR operation to remove the lung segmentation result at the EE phase from the lung segmentation result at the EI phase to obtain the lung excursion region segmentation result. Recall that our goal is to separate the hemi-chest wall excursion component and the hemi-diaphragm excursion component from the lung excursion region. This separation cannot be accomplished via simple morphological operations or connectivity analysis carried out on the lung excursion region segmentation due to the complex shape of lung excursion across different subjects. To accomplish the separation, our approach is to segment the hemi-diaphragm excursion region first and then to subtract this from the lung excursion region to obtain the hemi-chest wall component.

**Figure 6.**
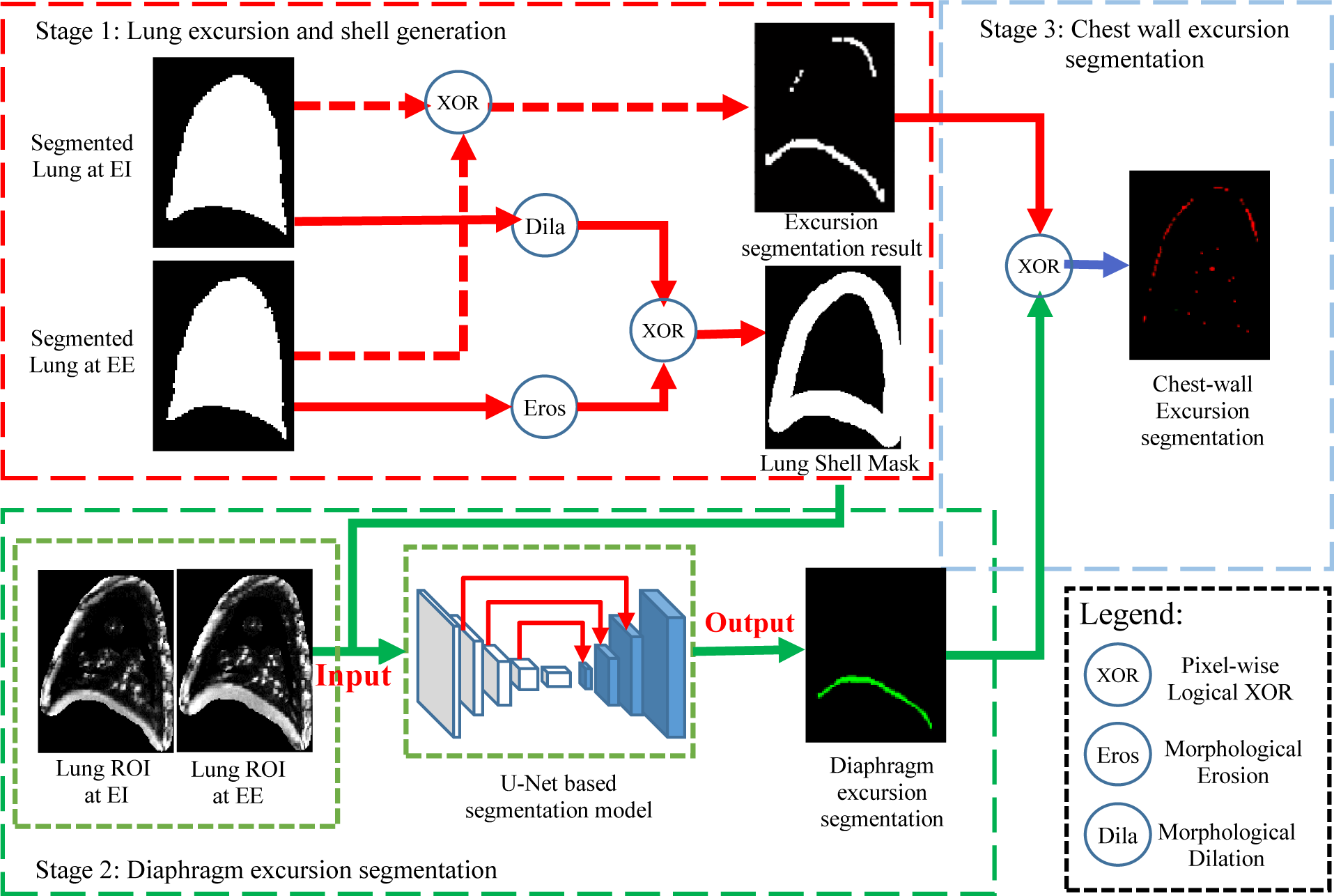
Illustration of shape-guided U-Net for lung excursion segmentation in dynamic MRI.

In the first stage, we computed the lung shell mask, which contains the distribution and shape information of lung excursion, using the following steps: i) morphological dilation of the lung segmentation result at the EI phase using a diamond-shaped structuring element of kernel size 5; ii) morphological erosion of the lung segmentation result at the EE phase using a diamond-shaped structuring element of kernel size 5; and iii) pixel-wise logical XOR operation between the erosion result and the dilation result. In the second stage, we computed the product between the lung segmentation masks and the MRI images to extract lung-shaped ROIs at the EI and EE phases. To ensure that the ROIs can cover the lung tissue, we utilized the dilation operation mentioned above. Then, we inputted the lung ROIs at EI and EI phases and the lung shell mask into a U-Net based segmentation network to predict the pixel-wise probability of the diaphragm excursion region. (The structure and loss function of lung excursion region segmentation network is the same as those employed for the lung tissue segmentation network.) Subsequently, we transformed the probability map into the binary diaphragm segmentation result by thresholding method with a threshold of 0.5. In the third stage, we calculated the chest wall excursion segmentation result by removing the diaphragm excursion region from the lung excursion result using pixel-wise logical XOR operation.

### 2.6 Post-processing

Due to the slender structure of the chest wall excursion component, the difficulty of clean chest wall excursion segmentation is greater than that of the diaphragm. The minor pixel-wise classification error in diaphragm excursion segmentation often leads to a high false positive rate in chest wall excursion segmentation. To handle this problem, we propose a post-processing method based on a convex hull algorithm and logical operations, as shown in Figure 7. Firstly, we compute the convex hull of the lung contour, which fills up the bottom of the lung segmentation. Then, we perform a logical subtraction operation and edge detection to calculate the bottom and contour regions of the convex hull, respectively. Next, the lung chest wall mask was computed by removing the bottom of the convex hull from the contour of the convex hull. Lastly, we integrated the chest wall mask with the chest wall excursion segmentation via pixel-wise logical AND operation.

**Figure 7.**
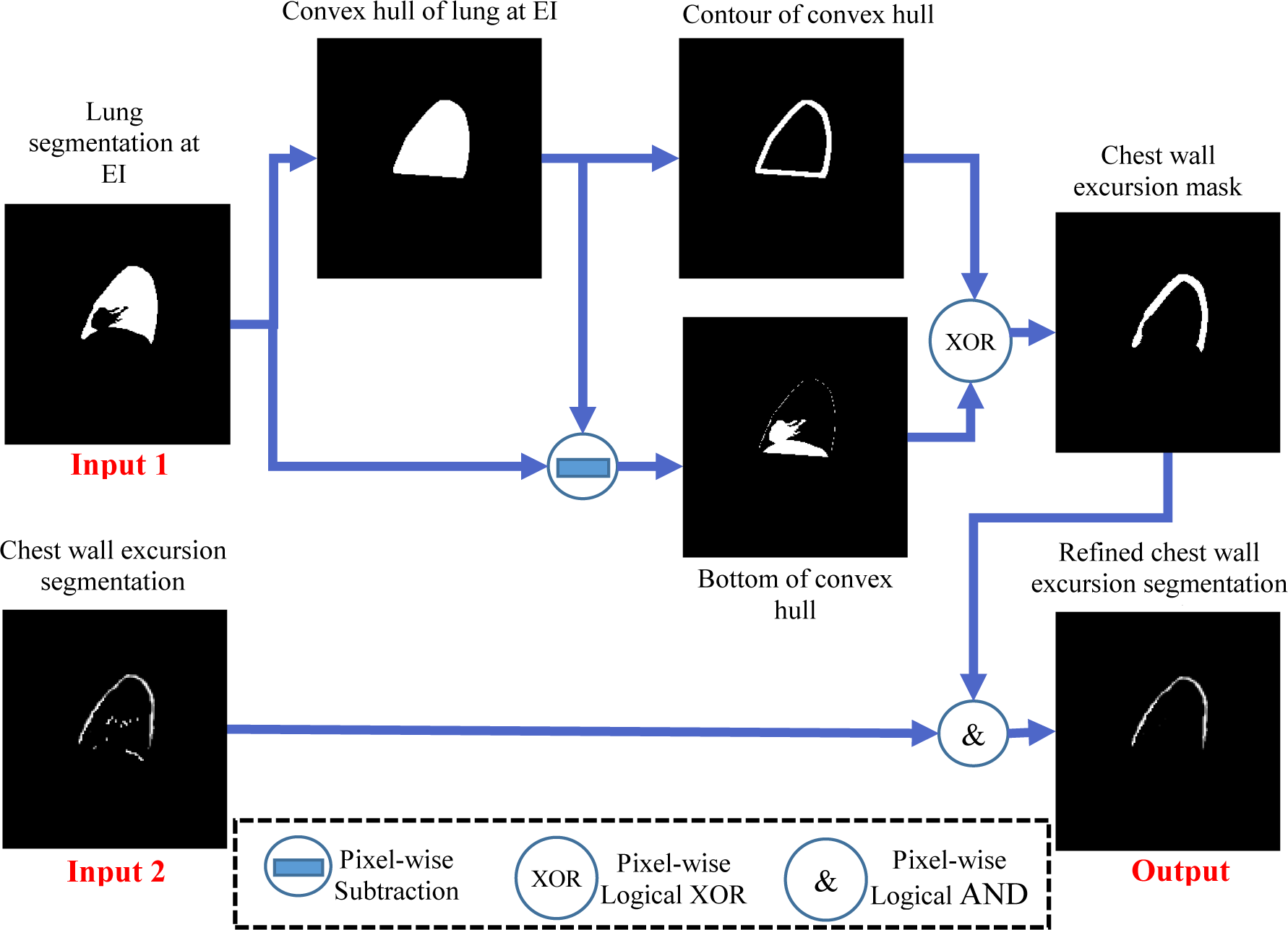
Illustration of post-processing for chest-wall excursion segmentation.

### 2.7 Data augmentation and DL model training

As is well known, the over-fitting problem frequently occurs in DL models due to insufficient training data. To handle this issue, we utilized two data augmentation methods to process each slice in dMRI: (1) resizing the image size with scale factors of 0.7, 0.9, 1.1, and 1.3 by a bilinear interpolation method for enriching the size diversity of lung, and (2) varying the intensity level of the image by multiplying the pixel-wise intensity value with scale factors of 0.9 and 1.1 for enriching the intensity diversity of lung.

At the training stage, we utilized the open-source DL platform TensorFlow [38] to implement the lung tissue and excursion segmentation networks. The trade-off parameters *λ*_1_and *λ*_2_in loss function were set to 0.5 and 10 based on initial experiments and subsequently fixed. We utilized random numbers under a normal distribution to initialize the lung tissue segmentation networks. Due to the strong relationship between lung tissue and excursion segmentation tasks, we utilized the parameters of the trained lung tissue segmentation networks to initialize the lung excursion networks. We employed Adam optimization to minimize the loss functions with the following hyper-parameters: learning rate 0.00001, batch size of training data 10, and number of iterations 1000. We computed the loss value on the validation data every 50 iterations and selected the network parameters with minimum loss values as the most suitable parameters.

## 3. Experiments & Evaluation

### 3.1 Experiments

We conducted a series of experiments on the testing data as summarized in Table 3. Experiments E1 to E5 were for investigating the effect of ROI strategy on lung segmentation performance, E6was to analyze the variability of lung volume estimation, E7-E8 were to compare the lung segmentation performance with the foundation models including Segment Anything Model (SAM) [39] and SAM-Med2D [40], and E9 to E12 were to assess excursion segmentation. Note that the SAM-Med2D model has a similar structure to SAM but was trained by a large medical dataset. The training and inference procedures in the experiments were as follows: (1) employing the same experimental environments and hyper-parameters to train the comparison models excepting that the batch size and number of iterations were set to 50 and 500, (2) utilizing the intensity standardization method and the min-max normalization model to pre-process the testing dMRI data, (3) inputting the pre-processed testing data into the trained lung tissue segmentation network to obtain the pixel-wise probability map, and transformed the probability map into the binary lung segmentation result by the thresholding method with a threshold value of 0.7, and (4) inputting the dMRI images and the lung tissue segmentation results on EI and EE into the trained lung excursion model to obtain the chest wall and diaphragm excursion volumes. Note that the iteration number of 500 can ensure the convergence of the loss function and reduce the training time.

**Table 3.**
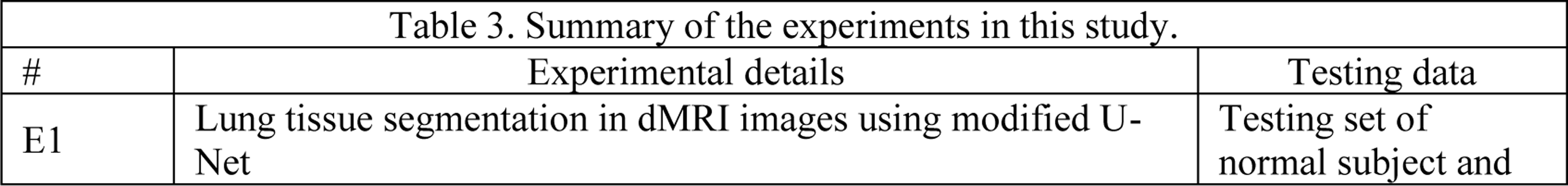
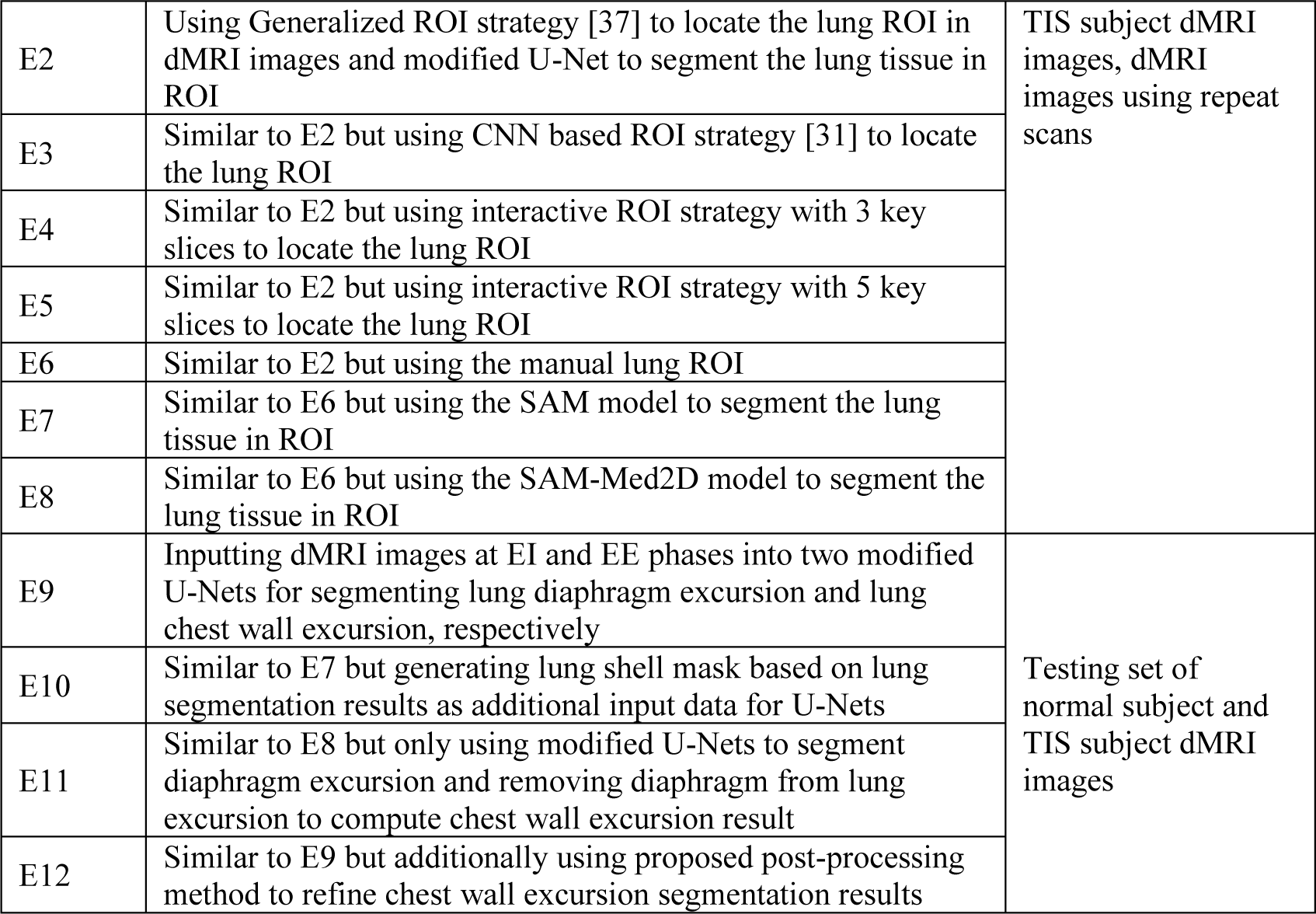
Summary of the experiments in this study.

### 3.2 Metrics

We applied multiple widely regarded metrics to quantitatively analyze lung tissue and lung excursion segmentation performance on testing data, including 3D Dice coefficient (DC) and max-mean Hausdorff distance (MM-HD) [37], defined as follows:

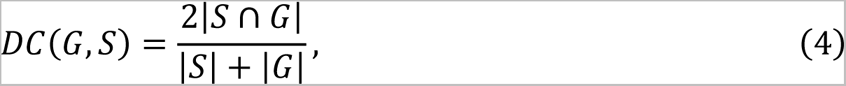

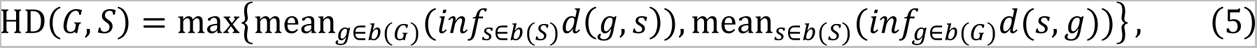

where *G* and *S* denote the set of voxels in binary ground truth and segmentation results, respectively, *b*(*G*) and *b*(*S*) denote the (2D) boundaries of *G* and *S* on slices, respectively, and *d*(*g*, *s*) indicates the (2D) Euclidean distance between *g* and *s*. Compared with the classical HD, the modified HD reduces the numerical sensitivity to small number of false positive voxels. Similarly, we utilized the 2D DC to evaluate the lung ROI accuracy for different ROI strategies, where *g* ∈ *G* and *s* ∈ *S* in Eq. 5 denote the pixels in the binary ground truth of bounding-boxes and the detected ROI results, respectively in the slice.

To assess the variability of our method on repeated dMRI scans, we designed a comparative test based on Coefficient of Variation (CV). Let the volume of the (left or right) lung under consideration in the 1^st^ scan be *v*_1_, *v*_2_, …, *v*_*n*_ and the volume in the 2^nd^ scan be *u*_1_, *u*_2_, …, *u*_*n*_ for the n cases. Then we find the coefficient of variation *CVi* for each sample *i* by

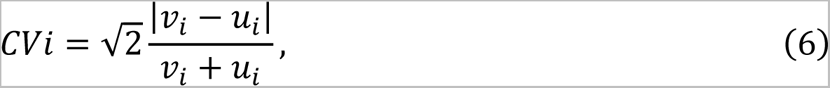

and find the mean and standard deviation of *CVi* values over all subjects to find the CV of our volume estimation method.

Note that the CV value can be influenced by several parameters during each scanning such as how the patient is positioned, the slice plane orientation relative to the anatomy, difference in the way the body interacts with the magnetic field, etc.

### 3.3 Lung ROI detection

In Figure 8 and Table 4, we illustrate lung ROI detection strategies. The results indicate that the proposed interactive ROI strategy achieves good agreement with the manual ROI and outperforms other comparative methods. The detection results of the generalized ROI (GROI) [37] and automatic ROI methods are much greater or smaller than the manual ROIs, leading to over-segmentation or under-segmentation of lung tissue. In addition, we observe that the 2D DC for the interactive ROI strategy with 5 initial ROIs is 0.91 and 0.90 for the normal and TIS testing data, respectively, which is 0.5 higher than those from the approach with 3 initial ROIs. Though more initial ROIs would improve lung detection accuracy, the cost of the interaction operation would increase at the same time. Considering the efficiency and accuracy of lung ROI detection, we generally utilized 5 initial ROIs to locate the right and left lungs in dMRI.

**Figure 8.**
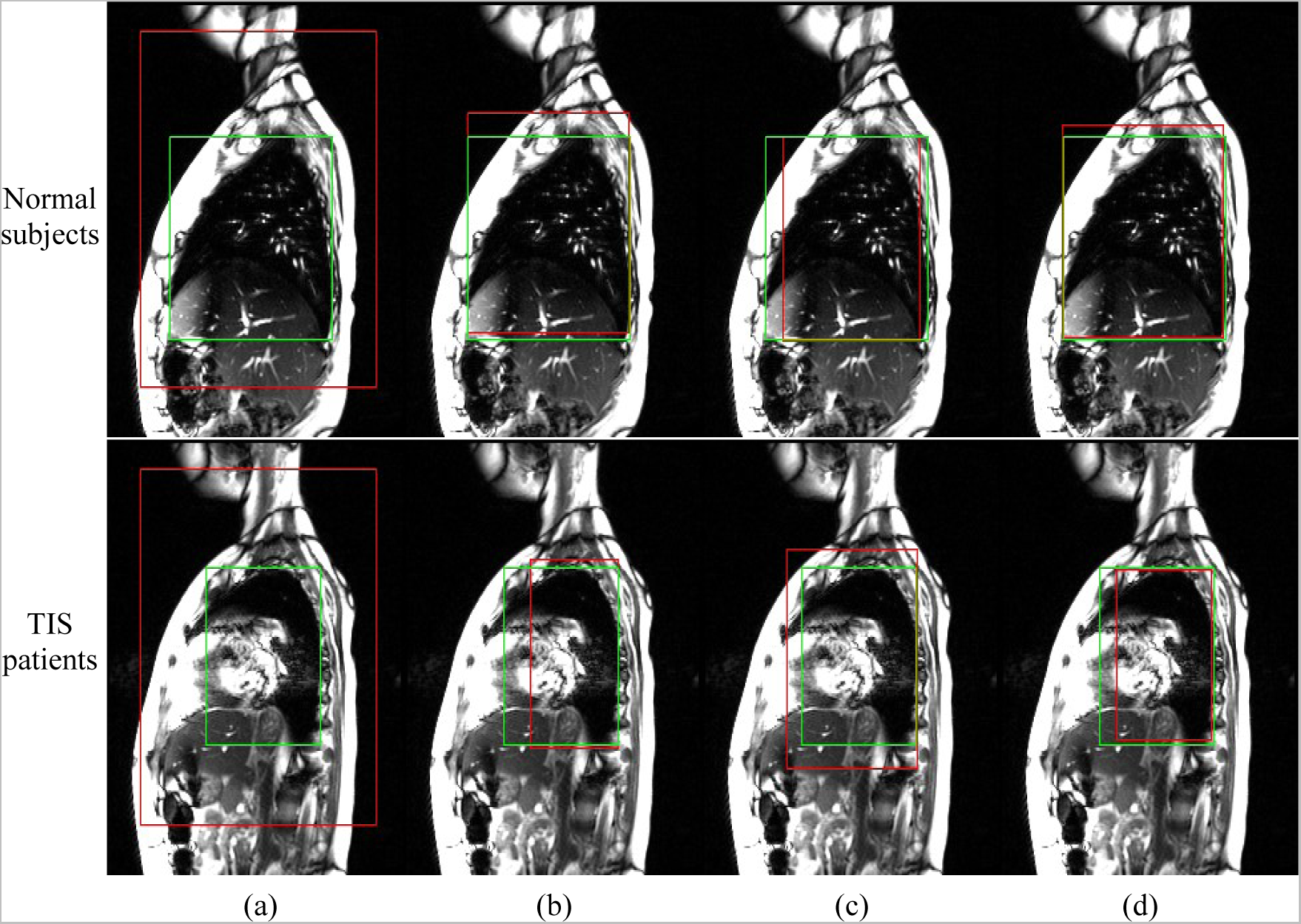
Examples of lung ROI detection in dMRI. The red and green rectangles represent automatic and manual ROIs, respectively. (a) Generalized ROI, (b) CNN based automatic ROI, (c) Interactive ROI with 3 initial ROIs, and (d) Interactive ROI with 5 initial ROIs.

**Table 4.** Comparison of quantitative results among ROI detection methods.

|  |  |  |  |
| --- | --- | --- | --- |
|  | ROI detection strategies | 2D DC |  |
|  |  | Normal subjects | TIS subjects |
| E2 | Strategy 1: Generalized ROI (GROI) | 0.23/0.12 | 0.32/0.18 |
| E3 | Strategy 2: CNN based automatic ROI | 0.92/0.13 | 0.84/0.21 |
| E4 | Strategy 3: Interactive ROI with 3 initial ROIs | 0.86/0.06 | 0.85/0.15 |
| E5 | Strategy 4: Interactive ROI with 5 initial ROIs | 0.91/0.14 | 0.90/0.16 |

### 3.4 Lung tissue segmentation

In Figure 9, we show four representative examples of lung tissue segmentation using 8 comparative experiments. We observe that most of the lung segmentation results have high similarity with the ground truth, especially for normal subjects. Due to the complex shape of the lungs in dMRIs of patients, the lung segmentation performance is poorer for TIS subjects. Also, the false-positive problem in the results of E1 and E2 is more obvious than in others, due to the interference from the surrounding tissue. The comparison of performance between E1 and other experiments demonstrates that the ROI detection strategies can efficiently improve lung segmentation accuracy. The comparison between E7 and E8 indicates that the large medical image dataset can enhance the lung segmentation performance of the foundation model.

**Figure 9.**
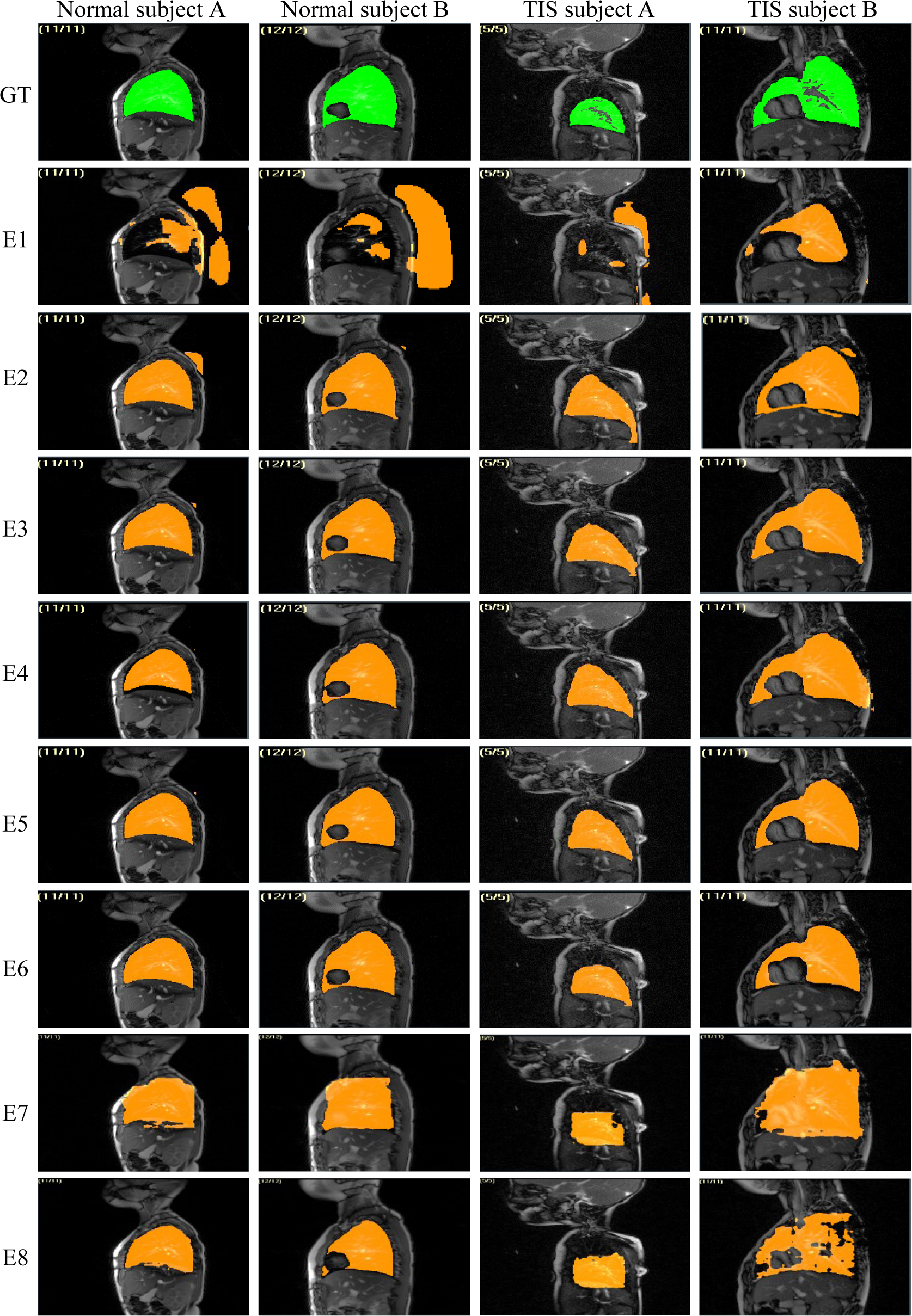
Examples of lung tissue segmentation in dMRI. The orange and green colors represent automatic and manual segmentations, respectively. 1^st^ row: Ground truth of lung segmentation. 2^nd^ – 9^th^: Automatic lung segmentation results from experiments E1-E8.

Table 5 summarizes the quantitative results of lung tissue segmentation for the 8 experiments. We observe that the model in E5 obtains high mean values of 0.96 and 0.89 and low standard deviations of 0.02 and 0.05 for 3D DC on normal and TIS subjects, respectively, indicating excellent lung segmentation accuracy and stability. The difference between E5 and E6 is very small, demonstrating that the accuracy of the lung ROI detection strategy is close to the pure manual approach. In contrast, there exists a significant difference of 3D DC value between experiments E1 and E2 and other experiments with p-values less than 0.05 using t-test, indicating that accurate lung ROI can significantly increase lung tissue segmentation performance for U-Nets. Furthermore, the proposed approach in experiment E5 achieves a low mean value of 3.58 and a low standard deviation of 3.27 for CV% on the repeat scan data, indicating high reproducibility. The proposed model in E6 outperforms the SAM-Med2D in E8 in terms of lung segmentation, especially on the TIS subjects, demonstrating that the foundation models, not refined by the corresponding medical image dataset, may exhibit inferior performance compared to the classic DL models.

**Table 5.** Lung segmentation on testing data from Normal and TIS data sets. All values are expressed as mean/standard deviation.

|  |  |  |  |  |  |  |
| --- | --- | --- | --- | --- | --- | --- |
|  | Lung segmentation methods | Normal subject testing data |  | Repeat scan data | TIS subject testing data |  |
|  |  | 3D DC | 2D MM-HD (mm) | CV% | 3D DC | 2D MM-HD (mm) |
| E1 | U-Net | 0.83/0.13 | 6.10/3.71 | 8.35/3.53 | 0.77/0.16 | 7.87/5.54 |
| E2 | Generalized ROI + U-Net | 0.84/0.14 | 8.23/7.56 | 7.75/3.62 | 0.80/0.13 | 8.26/4.52 |
| E3 | CNN based automatic ROI + U-Net | 0.96/0.03 | 2.13/7.14 | 4.12/4.2 | 0.82/0.10 | 6.87/5.08 |
| E4 | Interactive ROI (3 key slices) + U-Net | 0.93/0.04 | 3.46/2.54 | 5.84/2.49 | 0.85/0.07 | 5.25/3.70 |
| E5 | Interactive ROI (5 key slices) + U-Net | 0.96/0.02 | 2.73/1.36 | 3.58/3.27 | 0.89/0.05 | 4.49/3.95 |
| E6 | Manual ROI + U-Net | 0.98/0.01 | 1.90/0.76 | 2.95/3.03 | 0.90/0.05 | 4.08/2.72 |
| E7 | Manual ROI + SAM | 0.75/0.15 | 8.94/5.06 | 10.37/5.91 | 0.68/0.14 | 9.21/6.27 |
| E8 | Manual ROI + SAM-Med2D | 0.85/0.08 | 7.18/6.88 | 7.63/3.84 | 0.74/0.12 | 8.43/5.25 |

To demonstrate the effectiveness of 3D lung segmentation of our approach, we used CAVASS to display the manual and automatic segmentation results of the dMRIs as 3D surface renderings. Figure 10 illustrates four representative 3D visualization examples for 3 normal and 3 TIS subjects. We observe that the 3D lung segmentations show good agreement with the GT 3-dimensionally, preserving the complete anatomical structure of the right and left lungs with detail. The above results demonstrate the validity and robustness of our lung segmentation system.

**Figure 10.**
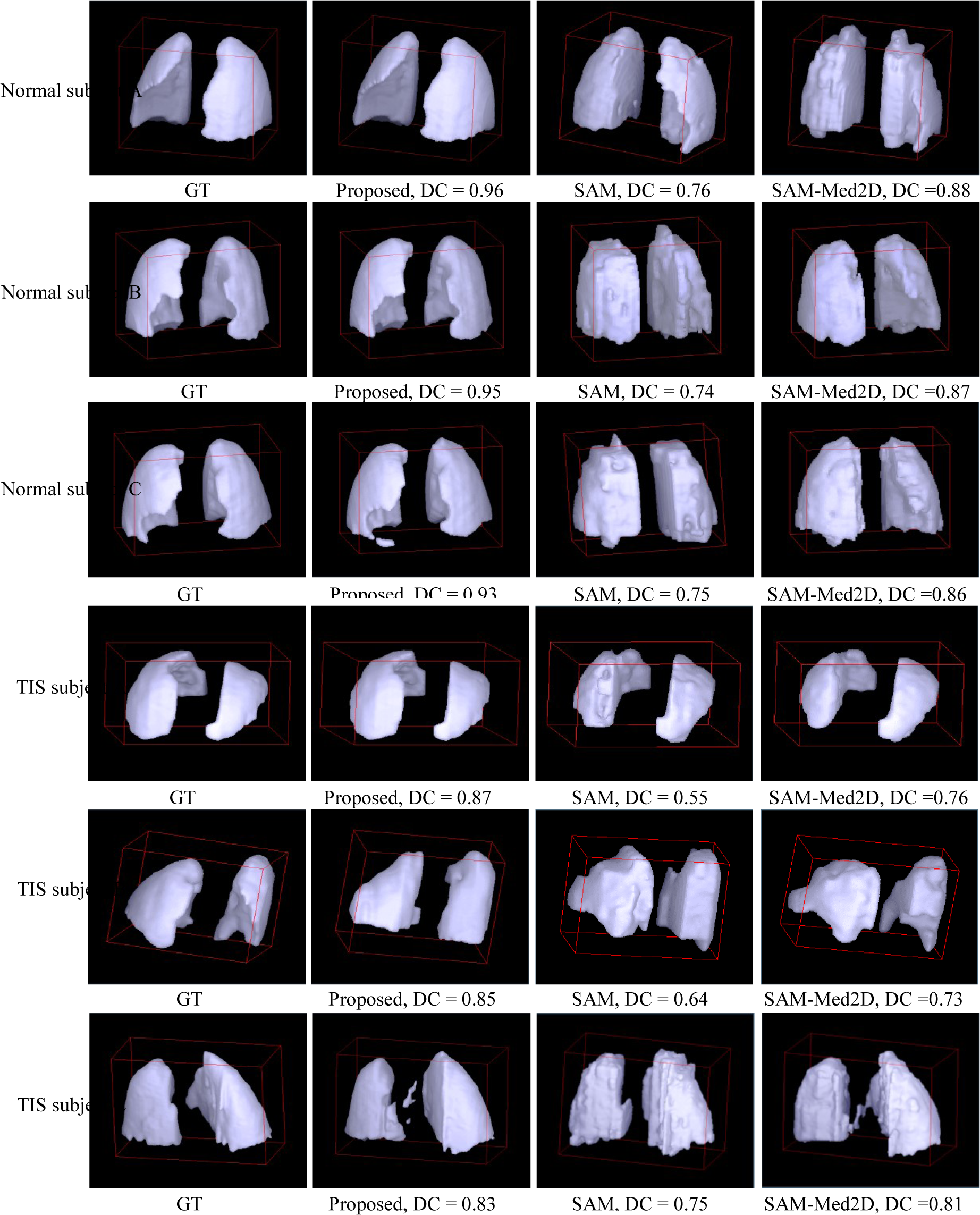
Illustration of lung 3D segmentation results: 1^st^-3^rd^ rows: From dMRI scans of 3 normal subjects. 4^th^-6^th^ rows: From dMRI scans of 3 TIS subjects. 1^st^ column: GT segmentations. 2^nd^ - 4^th^ columns: Automatic segmentations using proposed method, SAM, and SAM-2Med2D, respectively.

### 3.5 Lung excursion segmentation

Figure 11 shows 6 representative lung excursion segmentations from normal and TIS subjects, illustrating consistence with manual segmentations. We observe that the diaphragm excursion segmentation (shown in green) is better than that of the chest wall excursion (shown in red) for all experiments, due to the complex slender structure of the chest wall excursion component. By comparing the lung excursion segmentations of E11 and E12, we find that the proposed post-processing method can effectively reduce the false-positive pixels in the chest wall excursion segmentations.

**Figure 11.**
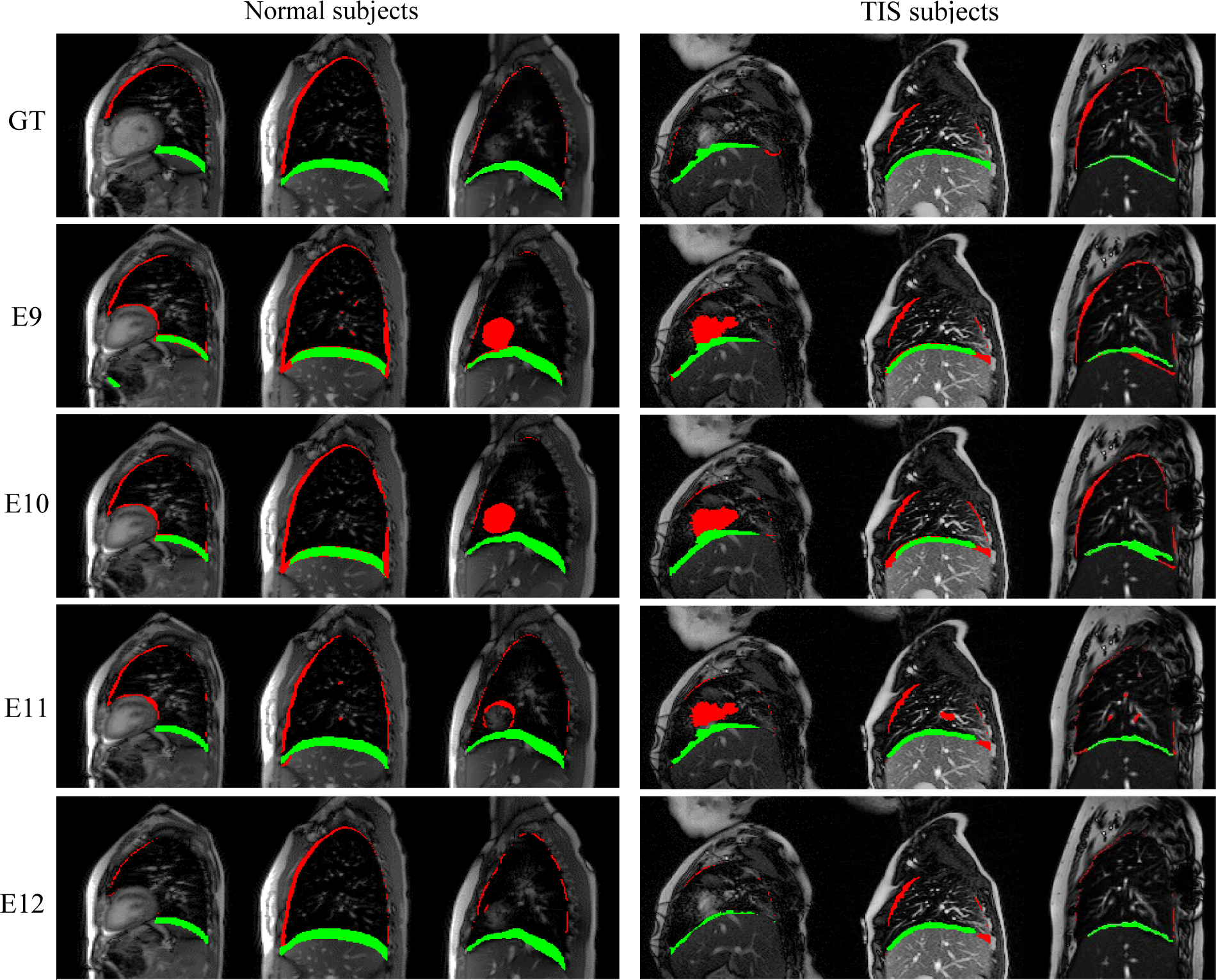
Examples of lung excursion segmentation in dMRI. The red and green colors represent chest wall and diaphragm excursion segmentations, respectively. 1^st^ row: Ground truth of excursion segmentation. 2^nd-^ 5^th^ row: Automatic lung excursion segmentation results from experiments E7-E10. 1^st^-3^rd^ columns: From normal subjects. 4^th^-6^th^. From TIS subjects.

Table 6 illustrates the quantitative results for experiments E9-E12. The proposed method achieves acceptably high mean values and low standard deviations for 3D DC on normal and TIS subjects. Due to the slender size and complex shape of the diaphragm and particularly chest wall excursion components, the Dice values can be considered as representing very good segmentations in view of the non-linear behavior of Dice for sparse objects compared to large non-sparse objects [41]. The comparison of results from experiments E9 and E10 demonstrates that the shape feature of lung shell can improve the diaphragm excursion segmentation. Also, we observe that the chest wall excursion segmentation performance of E11 is better than that of E10, illustrating that the chest wall excursion region produced by subtracting the diaphragm excursion from the lung tidal volume is more accurate than the chest wall excursion region predicted directly via U-Net.

**Table 6.** Diaphragm and chest wall excursion segmentations from testing data set of normal and TIS data sets. All values are expressed as mean/standard deviation.

|  |  |  |  |  |  |
| --- | --- | --- | --- | --- | --- |
|  | Lung excursion segmentation methods | 3D DC on Normal subject testing data |  | 3D DC on TIS subject testing data |  |
|  |  | Chest wall | Diaphragm | Chest wall | Diaphragm |
| E9 | Two U-Nets | 0.22/0.07 | 0.68/0.07 | 0.18/0.12 | 0.54/0.09 |
| E10 | Two U-Nets + Lung shell feature | 0.28/0.06 | 0.70/0.08 | 0.23/0.09 | 0.59/0.08 |
| E11 | Proposed shape-guided lung excursion segmentation model | 0.60/0.08 | 0.78/0.06 | 0.48/0.13 | 0.69/0.08 |
| E12 | Proposed shape-guided lung excursion segmentation model + Post-processing | 0.62/0.07 | 0.78/0.06 | 0.53/0.10 | 0.69/0.08 |

### 3.6 Tidal volume computation

In order to assess the clinical utility of our system, we computed the volumes of the left and right lungs at both EI and EE using both the proposed method and manual segmentations of the lung tissue, as displayed in Table 7. Additionally, we computed the coefficients of variation (CVs) and p-values to compare the automatically and manually generated measurements of the lungs. Our analysis reveals that our proposed approach yields a low mean value and a low standard deviation for CV, indicating a strong agreement between the two measurement methods. Furthermore, the majority of p-values exceed 0.05, demonstrating that the disparity between the proposed and manual measurements is not statistically significant.

**Table 7.** Lung volumes calculated on testing dataset of normal and TIS datasets. All values are expressed as mean/standard deviation.

|  |  |  |  |  |  |  |  |  |
| --- | --- | --- | --- | --- | --- | --- | --- | --- |
| Parameters (mL) | Normal subjects |  |  |  | TIS subjects |  |  |  |
|  | Manual | Proposed method | CV% | P-value | Manual | Proposed method | CV% | P-value |
| Left lung volume at end-inspiration | 527.82±196.83 | 464.97±170.7 | 8.88±2.51 | 0.091 | 250.98±180.98 | 209.36±163.62 | 22.05±23 | 0.124 |
| Right lung volume at end-inspiration | 690.16±228.97 | 636.28±209.01 | 5.83±2.44 | 0.222 | 371.03±244.7 | 313.61±236.62 | 17.07±15.64 | 0.129 |
| Left lung volume at end-expiration | 482.56±185.93 | 431.14±164.92 | 7.91±2.66 | 0.147 | 235.97±186.88 | 199.11±160.26 | 12.58±7.63 | 0.177 |
| Right lung volume at end-expiration | 633.82±218.13 | 589.93±201.32 | 5.14±2.2 | 0.298 | 336.7±238.07 | 295.63±232.59 | 20.58±26.5 | 0.265 |

We employed a linear regression plot to evaluate the concordance between manual and automatic measurements, as depicted in Figure 12. The regression coefficient for the measures exceeded 0.97 (range: R = 0.97 to 0.997), indicating a strong agreement with the ground truth.

**Figure 12.**
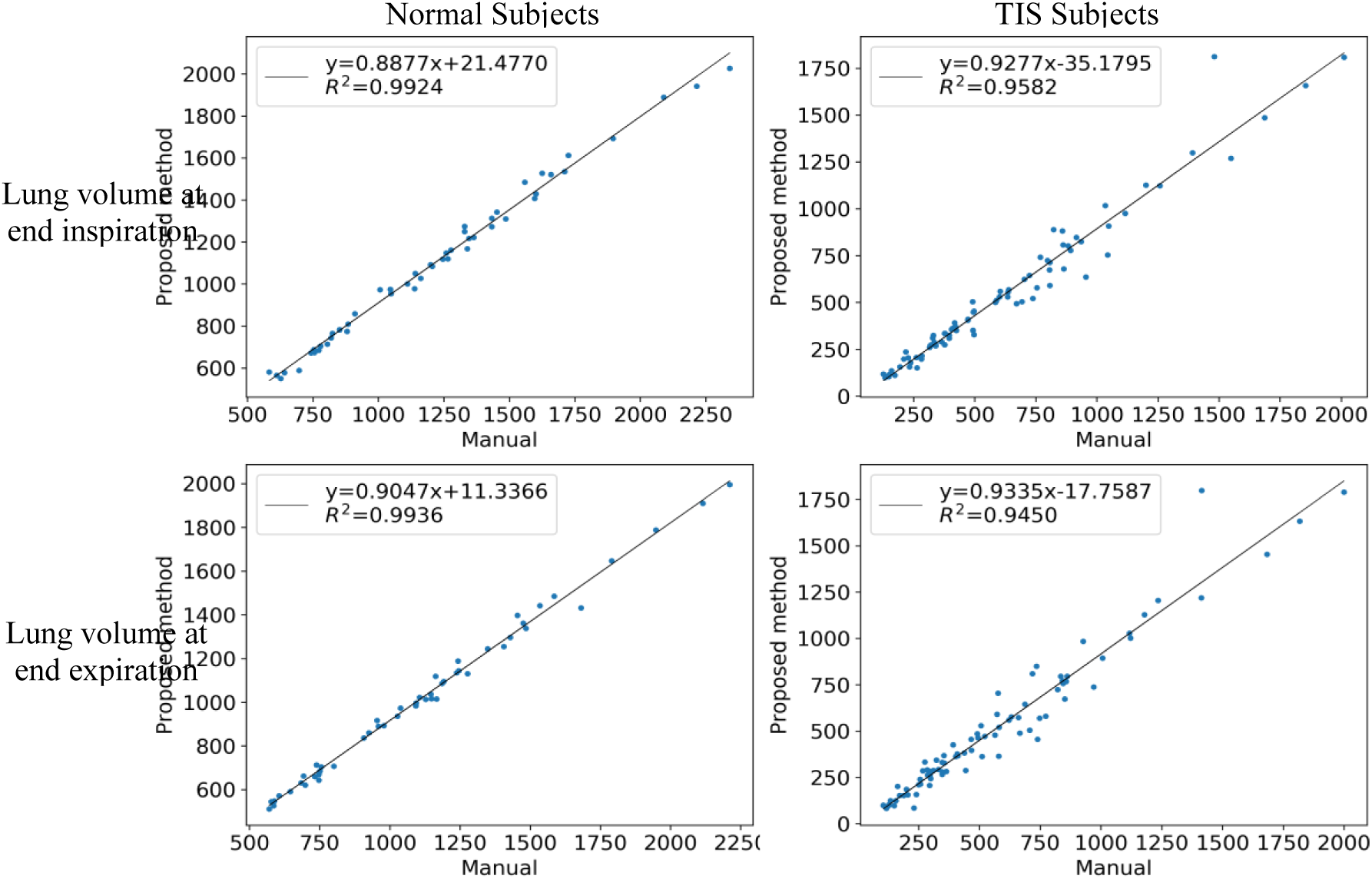
Linear regression plots of lung volume derived from dMRI scans using testing data. 1^st^ row: Lung volume at end-inspiration. 2^nd^ row: Lung volume at end-expiration. 1^st^ column: From normal subjects. 2^nd^ column: From TIS subjects.

### 3.7 Computational considerations

All experiments were conducted on a PC with an Intel i7-7700K CPU and two NVIDIA 1080 Ti GPUs. To assess the efficiency of our system, we computed the average processing time per subject and slice for each procedure, as shown in Table 8, which are found to be lower than 82 and 3 seconds, respectively. Note that the average processing time include the average time of 78.62 for interactive ROI strategy (5 key slices), which can save 44% time of manual ROI setting. The efficiency of our approach can meet the practical needs of image analysis of TIS patients for clinical or research purposes.

**Table 8.** Average processing time in seconds (per subject and per slice) for each procedure in our approach.

|  |  |  |  |  |
| --- | --- | --- | --- | --- |
| Methods | Proposed |  | SAM-Med2D |  |
| Procedures | Per subject | Per slice | Per subject | Per slice |
| Interactive operations | 78.62±5.97 | 2.03 | 141.8±13.49 | 3.68 |
| Pre-processing method for dMRI | 0.05±0.01 | < 0.01 | - | - |
| ROI-guided lung tissue segmentation model | 1.64±0.15 | 0.04 | 2.16±0.18 | 0.05 |
| Shape-guided lung excursion segmentation model | 1.27±0.06 | 0.03 | - | - |
| Post-processing method for chest wall segmentation result | 0.13±0.01 | < 0.01 | - | - |
| Total | 81.71±5.25 | 2.11 | 143.96±12.87 | 3.73 |

### 3.8 Comparison with published literature

As illustrated in Table 9, we compare the lung segmentation performance of our approach with several state-of-the-art methods. Our research work has several advantages over published studies as outlined below:

i) Data size: In our work, we acquired and annotated data sets from 101 normal subjects and TIS subjects with 248 dMRI scans as the experimental data, which are greater than those utilized in most other studies. We randomly selected 120 dMRI scans from the normal and TIS subjects as the testing data. In comparison, the study [29] utilized 669 and 74 multi-nuclear hyperpolarized gas MRI scans as the training data/testing data. It is known that less testing data would reduce the confidence of performance evaluation for DL-based models. Moreover, the gas MRI scans in that study were not dynamic scans as in our case, and hence the study cannot be strictly compared with our study.
ii) Interactive ROI strategy: This study provides an accurate, simple, and easy-to-use ROI extraction approach for dMRI scans, enabling good compatibility and generalizability of the approach to other lung segmentation applications. In addition, we conducted ablation experiments to demonstrate that the lung ROIs can effectively improve lung segmentation accuracy.
iii) Lung excursion segmentation: Distinct from other studies, we designed an efficient segmentation model to automatically delineate the chest wall and diaphragm excursion components of the lungs based on CNN and lung shell feature, contributing to calculation of important clinical measurements of lung structure and function for TIS patient assessment. Due to the complex shape of the chest wall excursion component, their false-positive and false-negative challenges are more serious than for the diaphragm excursion component. To improve the segmentation performance for the chest wall component, we proposed a post-processing method to refine the results.
iv) TIS application: The acquired dMRI data set includes 46 subjects with TIS. The proposed method achieves good segmentation performance for both normal subjects and TIS subjects. In contrast, other studies did not evaluate the lung tissue segmentation performance of subjects with TIS where the anatomy undergoes severe distortions. Also, this study assessed lung tidal volume, chest wall excursion volume, and diaphragm excursion volume results, which are clinically relevant measurements in patients with TIS and other respiratory diseases. The experimental data shown in Table 7 and Figure 12 demonstrate that the automatic estimation of the clinically relevant measurements has good agreement with that derived from GT segmentations. Therefore, our approach is amenable for routine use in patients.
v) Segmentation performance: Due to the lack of other publicly available thoracic dynamic MRI datasets, we cannot conduct a direct comparison of performance with other methods published in the literature. The main factors potentially influencing the lung segmentation performance include the size of the testing data, the image quality of the testing data, the quality of manual annotation, etc. We utilized the largest possible independent data set to estimate the metric values of performance for our approach. The mean value of DC based on our approach is competitive with that of other studies, with a standard deviation lower than that of most other methods. We randomly selected 100/80 3D dMRI frames from normal subjects as the training/testing data and achieved a mean value of 0.96 for 3D DC. In comparison, the study [29] obtained a mean value of 0.96 using 669 and 74 MRI scans as the training data/testing data. Compared with the SAM-Med2D model, which was trained by millions of medical images, our method also has higher efficiency and accuracy in terms of lung segmentation.

**Table 9.** Comparison with lung segmentation methods in the literature.

3.8 Comparison with published literature
| Table 9. Comparison with lung segmentation methods in the literature. |  |  |  |  |  |
| --- | --- | --- | --- | --- | --- |
| Reference | Year | Approach | Data type | Number of training/testing cases | Metrics |
| Mansoor et al. [19] | 2014 | Fuzzy connectedness image segmentation algorithm | Chest CT | 0/403 scans | DC=0.96;<br>HD=18.82 mm; |
| Tustison et al. [17] | 2016 | Atlas-based | Multi-sequence Proton MRI | 0/62 scans | DC = 0.98 ± 0.01 |
| Tong et al. [20] | 2017 | Interactive iterative relative fuzzy connectedness image segmentation algorithm | Thoracic dynamic MRI | 0/39 scans | True Positive Rate = 0.91±0.03;<br>HD = 2.15±0.33 mm; |
| Hu et al. [27] | 2020 | Mask R-CNN | Chest CT | 998/267 slices | DC = 0.97±0.03 |
| Chen et al. [42] | 2020 | 3D FCN | Dual-energy CT | 27/9 scans | DC = 0.98±0.02 |
| Willers et al. [22] | 2020 | Artificial Neural Network | Matrix pencil decomposition MRI | 51/25 scans | DC = 0.94 ± 0.01 |
| Astley et al. [29] | 2020 | 3D CNN | Multi-nuclear hyperpolarized gas MRI | 669/74 scans | DC = 0.96± 0.02;<br>HD = 2.22 ± 2.16 mm; |
| Zhao et al. [28] | 2021 | 3D V-Net | Chest CT | 90/22 scans | DC = 0.98 |
| Astley et al. [30] | 2023 | DL-based | Multi-sequence proton MRI | 593/82 scans | DC = 0.96;<br>HD = 1.63 mm; |
| Cheng et al. [40] | 2023 | Manual ROI and SAM-Med2D model | Thoracic dynamic MRI | 4.6M images and 19.7 masks/Unknown | i) Normal subjects: 3D DC = 0.85±0.08, 2D MM-HD = 7.18/6.88 mm;<br>ii) TIS patients: 3D DC = 0.74/0.12, 2D MM-HD = 8.43/5.25 mm; |
| Proposed | 2023 | Interactive ROI strategy and ROI-guided CNN | Thoracic dynamic MRI | i) Normal subjects: 100/80 scans;<br>ii) TIS subjects: 44/40 scans; | i) Normal subjects: 3D DC = 0.96±0.02, 2D MM-HD = 2.73/1.36 mm;<br>ii) TIS patients: 3D DC = 0.89/0.05, 2D MM-HD = 4.49/3.95 mm; |

### 3.9 Limitations

There are some limitations in our method. Firstly, the lung segmentation system for dMRI requires users to set 10 corner-points of the initial ROI for the right and left lungs separately. We are working on simplifying this interactive operation. Second, the lung segmentation performance of our method heavily depends on the lung ROI detection result. For some lungs with extreme distortions as encountered in some TIS patients, it may be very difficult to recognize the location of the lungs, even for experts, resulting in disconnected lungs in the segmentation result. Third, the lung excursion segmentation accuracy of our system can be improved. We are designing a novel segmentation network by integrating more TIS-specific hand-crafted features with deep learning features to enhance the chest wall excursion segmentation performance.

## 4. Conclusions

In this paper, we utilized deep learning techniques to construct a semi-automatic system for lung tissue and lung excursion segmentation in thoracic dynamic magnetic resonance imaging for the quantitative analysis of lung function. Our method can be divided into three stages: 1) extraction of the lung ROI from dMRI slices using the interactive ROI strategy for reducing the negative influence of the tissue located far away from the lung, 2) segmentation of the lung tissue in the lung ROIs using a modified 2D U-Net, and 3) segmentation of the lung excursion region in the lung ROIs using the lung shell based on 2D U-Net. The quantitative and illustrative experimental results performed on a large and independent testing data set demonstrate that our approach achieves excellent agreement with manual detection and segmentation of lung tissue and lung excursion region in dMRI both in normal subjects and TIS patients. In addition, the computation of lung tidal volumes using our system is close to that derived manually, indicating strong potential for clinical utility in the assessment of patients with TIS.

## Data Availability

All data produced in the present study are available upon reasonable request to the authors

## Acknowledgments

This work is supported by an NIH grant HL130468.

